# SARS-CoV-2 triggered neutrophil extracellular traps (NETs) mediate COVID-19 pathology

**DOI:** 10.1101/2020.06.08.20125823

**Authors:** Flavio Veras, Marjorie Pontelli, Camila Silva, Juliana Toller-Kawahisa, Mikhael de Lima, Daniele Nascimento, Ayda Schneider, Diego Caetité, Roberta Rosales, David Colón, Ronaldo Martins, Italo Castro, Glaucia Almeida, Maria Isabel Lopes, Maíra Benatti, Letícia Bonjorno, Marcela Giannini, Rodrigo Luppino-Assad, Sérgio de Almeida, Fernando Vilar, Rodrigo Santana, Valdes Bollela, Maria Martins, Carlos Miranda, Marcos Borges, Antônio Pazin-Filho, Larissa Cunha, Dario Zamboni, Felipe Dal-Pizzol, Luiz Leiria, Li Siyuan, Sabrina Batah, Alexandre Fabro, Thais Mauad, Marisa Dolhnikoff, Amaro. Duarte-Neto, Paulo Saldiva, Thiago Cunha, José Alves-Filho, Eurico Arruda, Paulo Louzada-Junior, Renê de Oliveira, Fernando Cunha

## Abstract

Severe COVID-19 patients develop acute respiratory distress syndrome that may progress to respiratory failure. These patients also develop cytokine storm syndrome, and organ dysfunctions, which is a clinical picture that resembles sepsis. Considering that neutrophil extracellular traps (NETs) have been described as an important factors of tissue damage in sepsis, we investigated whether NETs would be produced in COVID-19 patients and participate in the lung tissue damage. A cohort of 32 hospitalized patients with a confirmed diagnosis of COVID-19 and respective healthy controls were enrolled. NETs concentration was assessed by MPO-DNA PicoGreen assay or by confocal immunofluorescence. The cytotoxic effect of SARS-CoV-2-induced NETs was analyzed in human epithelial lung cells (A549 cells). The concentration of NETs was augmented in plasma and tracheal aspirate from COVID-19 patients and their neutrophils spontaneously released higher levels of NETs. NETs were also found in the lung tissue specimens from autopsies of COVID-19 patients. Notably, viable SARS-CoV-2 can directly induce *in vitro* release of NETs by healthy neutrophils in a PAD-4-dependent manner. Finally, NETs released by SARS-CoV-2-activated neutrophils promote lung epithelial cell death *in vitro*. These results unravel a possible detrimental role of NETs in the pathophysiology of COVID-19. Therefore, the inhibition of NETs represent a potential therapeutic target for COVID-19.

## Introduction

The coronavirus disease 2019 (COVID-19) became pandemic, affecting more than four million people worldwide, with more than three hundred thousand deaths until May 2020. Caused by the severe acute respiratory syndrome coronavirus 2 (SARS-CoV-2), COVID-19 resembles influenza, with a clinical picture ranging from mild upper airway symptoms in the majority of cases to severe lower airway symptoms in a subgroup of patients, in which acute respiratory distress syndrome (ARDS) develops and may rapidly progress to respiratory failure due to intense acute lung injury, its major cause of death (Lai et al., 2020). It is also known that this subgroup of patients has a cytokine storm syndrome, which seems to be responsible for multi-organ failure (Chen et al., 2020). In addition, COVID-19 patients develop signs and symptoms similar to those observed in sepsis, much of which a result of microthrombosis, organ dysfunctions, and eventually shock (Wu and McGoogan, 2020; Magro et al., 2020; Guan et al., 2020).

The increased number of circulating neutrophils has been described as an indicator for the severity of respiratory symptoms and a poor clinical outcome in COVID-19 (Guan et al., 2020). Our group and others have described the pathogenic role for neutrophils and their neutrophil-derived extracellular traps (NETs) as a critical factor mediating tissue damage in vital organs during sepsis (Colón et al., 2019; Czaikoski et al., 2016; Kambas et al., 2012). NETs are networks of extracellular fibers composed of DNA containing histones and granule-derived enzymes, such as myeloperoxidase (MPO) and elastase (Brinkmann et al., 2004). The process of NETs formation by neutrophils, denominated NETosis, has been widely studied. In general, the process starts with neutrophil activation by pattern recognition receptors (PRR) or chemokines, followed by reactive oxygen species (ROS) production and calcium mobilization, which leads to the activation of protein arginine deiminase 4 (PAD-4), an intracellular enzyme involved in the deimination of arginine residues on histones (Li et al., 2010).

The NETosis results in the capture and killing of pathogens; however, increasing evidence has shown that NETs have also been implicated in tissue injury and, consequently, on the pathogenesis of several diseases (Brinkmann et al., 2004; Colón et al., 2019). We described that, during experimental and clinical sepsis, NETs are found in high concentrations in the blood and positively correlated with biomarkers of vital organ injuries and sepsis severity. Furthermore, disruption or inhibition of NETs release by pharmacological treatment with recombinant human DNase (rhDNase) or PAD-4 inhibitors, respectively, markedly reduced the organs damage, especially in the lungs, and increased the survival rate of severe septic mice (Colón et al., 2019).

Given the well-known association of NETosis with key events also involved in the COVID-19 pathophysiology, such as cytokine overproduction (Mehta et al., 2020), microthrombosis (Magro et al., 2020; Dolhnikoff et al., 2020), and ARDS (Lai et al., 2020), we hypothesized that NETs are triggered during SARS-CoV-2 infection and might contribute to tissue injury. On confirming this hypothesis, NETs could become potential targets for the treatment of COVID-19.

## Results and Discussion

### COVID-19 patients’ blood neutrophils produce higher levels of NETs

Although the number of patients with COVID-19 is growing exponentially worldwide, there is no effective treatment for this disease. The knowledge of the mechanisms by which the host deals with the SARS-CoV-2 infection will certainly allow the development of new therapeutic strategies aiming to treat COVID-19. Here, we are reporting that in severe COVID-19, circulating and lung-infiltrating neutrophils are releasing higher levels of NETs. We also showed that SARS-CoV-2 directly stimulates neutrophils to release NETs, and these activated neutrophils are highly cytotoxic for lung epithelial cells in a mechanism dependent on NETs, which was mimicked by purified NETs. Firstly, we confirmed the SARS-CoV-2 infection of all patients (32 patients) by RT-PCR of nasopharyngeal samples using standard protocol (Nalla et al., 2020) (**Supplementary Table 1**) and/or detection of specific antibodies IgM and IgG against SARS-CoV-2 in plasma samples. The demographic and clinical characteristics of COVID-19 patients are shown in **Supplementary Table 1**. The increased number of circulating neutrophils is an indicator of a worse outcome in COVID-19 (Huang et al., 2020; Chen et al., 2020). We evaluated the number of circulating neutrophils (CD15^+^CD16^+^ cells) in COVID-19 patients by flow cytometry. As recently reported (Magro et al., 2020), the COVID-19 patients of the present cohort also showed a significant neutrophilia (**Fig. S1**). Although, it has been described that COVID-19 patients also present a prominent influx of neutrophils into the lungs and, possibly, other vital organs, the pathologic role of neutrophils during COVID-19 has not been demonstrated yet (Dolhnikoff et al., 2020; Tian et al., 2020). In 2004, Brinkmann and collaborators initially described NETs as a microbicidal tool released by neutrophils (Brinkmann et al., 2004). However, accumulating evidence demonstrated that NETs have double-edged-sword activities. Besides their microbicidal activity, NETs are the cause of tissue damage in several inflammatory diseases, including rheumatoid arthritis (Khandpur et al., 2013; Sur Chowdhury et al., 2014), diabetes (Wong et al., 2015) and sepsis (Colón et al., 2019; Czaikoski et al., 2016; Kambas et al., 2012). The levels of NETs in the circulation and organ tissues are increased during sepsis, and the inhibition of NETs production prevents lungs and other organs lesions observed during experimental sepsis (Colón et al., 2019). Considering that several sepsis events, such as cytokine storm, vital organs lesions are also also observed in patients with severe COVID-19 (Chen et al., 2020; Guan et al., 2020), we then determined the concentration of soluble NETs in the plasma by measuring MPO-DNA complexes (Colón et al., 2019). Higher levels of NETs were found in the plasma of COVID-19 patients compared to healthy controls (**Fig. 1 A**).

**Figure 1.**
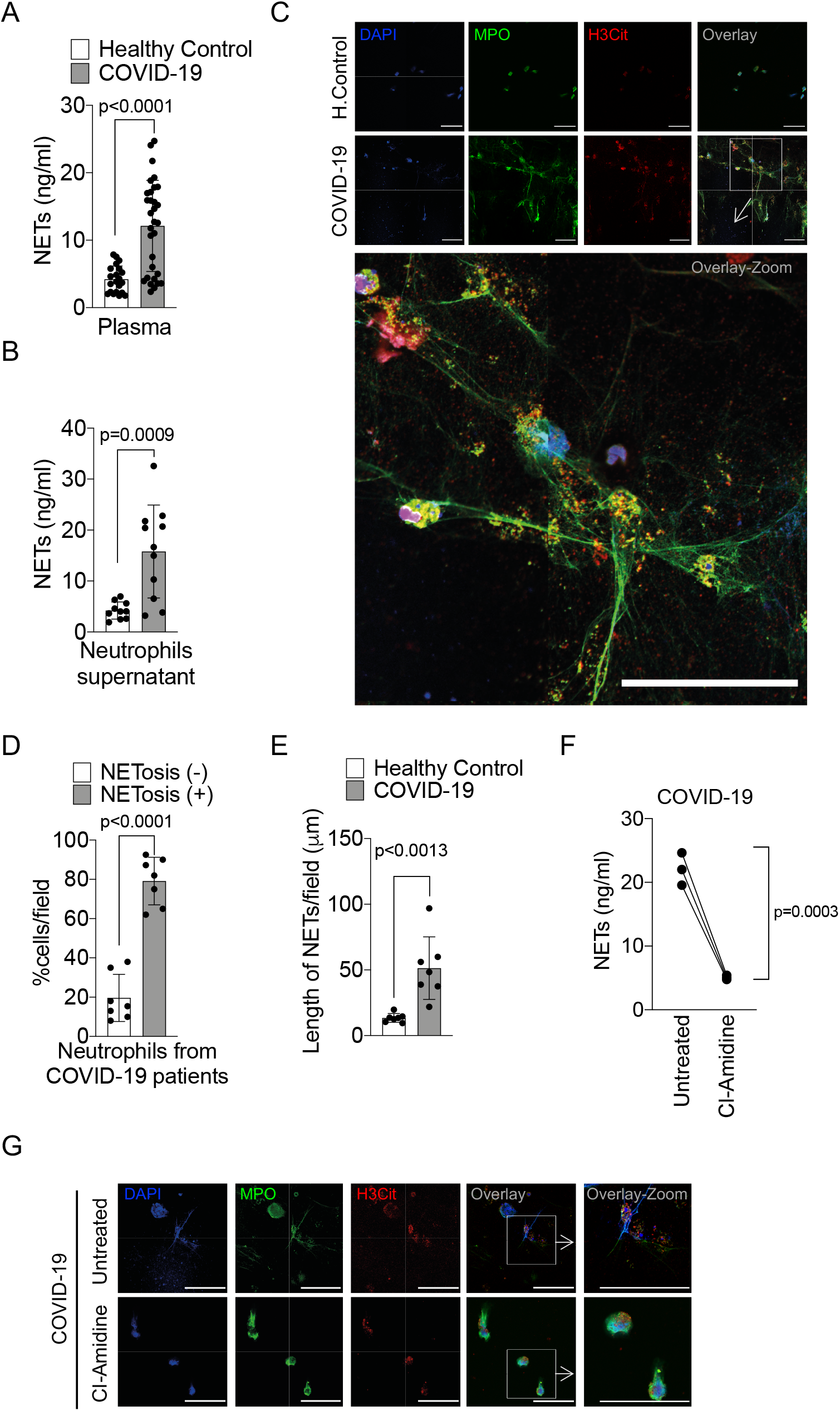
COVID-19 patients produce high concentrations of NETs. Plasma and neutrophils were isolated from healthy controls and COVID-19 patients. (A) NETs quantification by MPO-DNA PicoGreen assay in plasma from healthy controls (n=21) or COVID-19 patients (n=32). Data are the mean ± SD, two-tailed unpaired Student t-test. (B) Supernatants from cultures of blood isolated neutrophils from healthy controls (n=10) or COVID-19 patients (n=11). NETs quantification was performed using MPO-DNA PicoGreen assay. Data are the mean ± SD, two-tailed unpaired Student t-test. (C) Representative confocal analysis of NETs release by neutrophils isolated from healthy controls (n=10) or COVID-19 patients (n=11), cultured for 4 h at 37 °C. Cells were stained for nuclei (DAPI, blue), myeloperoxidase (MPO, green), and Citrullinated Histone H3 (H3Cit, red). Scale bar indicates 50 μm. (D) Percentage of NETosis in neutrophil from COVID-19 patients (n=7). Data are the mean ± SD of 5 fields/group, two-tailed unpaired Student t-test. (E) NETs length quantification. Data are the mean ± SD of 5 fields/group, two-tailed unpaired Student t-test. (F) NETs quantification by MPO-DNA PicoGreen assay in the supernatants of blood isolated neutrophils from COVID-19 patients (n=3) pre-incubated, or not, with PAD-4 inhibitor (Cl-Amidine; 200 μM) for 4h at 37°C. Data are the mean ± SD, two-tailed paired Student t-test. (G) Representative confocal images showing the presence of NETs in isolated neutrophils from COVID-19 patients, treated or not, with Cl-Amidine (200 μM). Cells were stained for nuclei (DAPI, blue), MPO (green), and H3Cit (red). Scale bar indicates 50 μm.

To verify whether the higher concentration of NETs in the plasma of COVID-19 patients was a consequence of the increased number of circulating neutrophils or their enhanced ability to release these mediators, we purified blood neutrophils from controls and COVID-19 patients to analyze the spontaneous release of NETs *in vitro*. Using a normalized number of cells, we found that blood neutrophils from COVID-19 patients released higher levels of NETs compared to controls (**Fig. 1 B**). NETs can be microscopically visualized as extracellular fibers of DNA colocalizing with MPO and citrullinated histone H3 (H3Cit) (Brinkmann et al., 2004). Confirming, confocal microscopy analysis also showed the increased ability of neutrophils from COVID-19 patients to release NETs spontaneously (**Fig. 1 C**). NETs from COVID-19 patients are typical rigid rods. Notably, more than 80% of neutrophils from COVID-19 patients were positive for the NETs structures (**Fig. 1 D**). In addition, the branch lengths of the spontaneously released NETs by neutrophils from COVID-19 patients were also larger than those from healthy controls (**Fig. 1 E**).

One of the most important intracellular mechanisms that mediate the release of NETs by neutrophils is the activation of PAD-4. This enzyme catalyzes the citrullination of arginine residues on histones to promote chromatin decondensation and extrusion of NETs (Li et al., 2010). Notably, the spontaneous release of NETs by blood neutrophils from COVID-19 patients was reduced by the incubation of cells with Cl-Amidine, an inhibitor of PAD-4 (**Fig. 1, F and G**). These results indicate that in COVID-19 patients, circulating neutrophils are more susceptible to the release of PAD4-dependent NETs, which might cause a systemic increase of soluble NETs. In agreement, during the preparation of this manuscript, a study was published supporting these findings (Zuo et al., 2020).

### NETs are highly detected in the tracheal aspirate and lung tissue from COVID-19 patients

Lung inflammation is the primary cause of the life-threatening respiratory disorder at the critical and severe forms of COVID-19 (Guan et al., 2020). NETs have been identified in the lung tissue of viral and non-viral infected patients and experimental animals (Sivanandham et al., 2018; Dicker et al., 2018). We next investigated the content of NETs in the tracheal aspirate obtained from patients with severe COVID-19 under mechanical ventilation admitted to the intensive care unit (ICU) or in the lung sections from *postmortem* COVID-19. Firstly, we found a higher concentration of NETs in the tracheal aspirate from COVID-19 patients compared to airway fluid from healthy controls (**Fig. 2 A**). The concentration of NETs in the tracheal aspirate was 10 times higher than observed in the plasma of COVID-19 patients (**Fig. 1 A and Fig. 2 A**). The confocal microscopy analysis of pellet cells from the tracheal aspirate also revealed typical NETs structures with extracellular DNA co-staining with MPO and H3Cit (**Fig. 2 B**).

**Figure 2.**
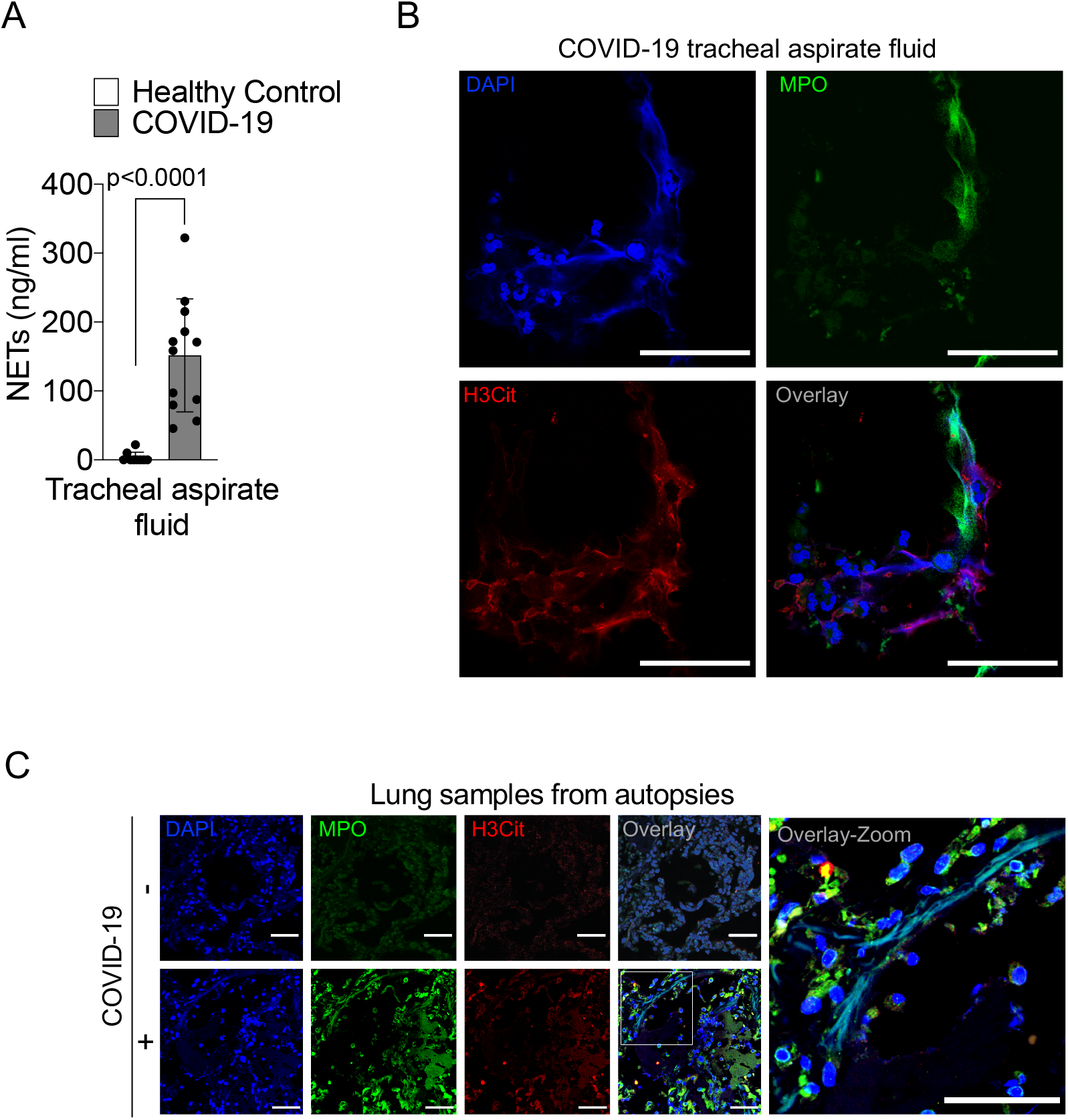
NETs release in the lungs of COVID-19 patients. (A) NETs quantification by MPO-DNA PicoGreen assay in the tracheal aspirate from COVID-19 patients (n=12) and in saline-induced airway fluids from healthy controls (n=9). Data are the mean ± SD, two-tailed unpaired Student t-test. (B) Representative confocal analysis of NETs in neutrophil from the tracheal aspirate of COVID-19 patients (n=5). Cells were stained for nuclei (DAPI, blue), MPO (green), and H3Cit (red). Scale bar indicates 50 μm. (C) Representative confocal images of the presence of NETs in the lung tissue from autopsies of negative controls (n=3) or COVID-19 (n=3) patients. Cells were stained for nuclei (DAPI, blue), MPO (green), H3Cit (red). Scale bar indicates 50 μm.

We next investigated whether lung tissue damage observed in COVID-19 patients was associated with the local presence of NETs. Firstly, we confirmed extensive injury in the lungs of COVID-19 patients by CT analysis (**Fig. S2 A**). We observed in CT imaging multiple consolidations with air bronchograms in all pulmonary fields, with peripheral and peribronchovascular distribution, more evident in the lower lobes, associated with ground-glass opacities, compatible with diffuse alveolar damage (**Fig. S2 A**). Next, we performed a minimally invasive autopsy to harvest lung tissue and analyzed histopathologic features. The histopathological analysis of the lung tissue from COVID-19 autopsies exhibited exudative/proliferative diffuse alveolar damage (**Fig. S2 B-I**); intense alveolar and small airway epithelial changes with cytopathic effects and squamous metaplasia (**Fig. S2 B-II**); mild lymphocytic infiltration; changes in pulmonary arterioles (endothelial swelling and small fibrinous thrombi). Neutrophilic pneumonia was observed in the samples from 6 out of 10 patients, in variable degrees (**Fig. S2 B**). The confocal microscopy analysis of these lung tissues from COVID-19 *postmortem* individuals also revealed the presence of neutrophil infiltration and characteristic NETs structures with extracellular DNA staining colocalizing with MPO and H3Cit (**Fig. 2 C**). NETs were not found in the healthy tissues obtained from lobectomy due to heart failure (**Fig. 2 C**). These results indicate that NETs are released in the lung tissue and are associated with lung damage in COVID-19 patients.

### SARS-CoV-2 replication induces the release of NETs

Several stimuli trigger NETosis, including pathogen-associated molecular patterns (PAMPs), damage-associated molecular patterns (DAMPs), and inflammatory mediators, including cytokines and chemokines (Brinkmann et al., 2004; Keshari et al., 2012). There is also evidence that infection of neutrophils with several viruses triggered the formation of NETs (Hiroki et al., 2020; Funchal et al., 2015; Saitoh et al., 2012), which led us to investigate whether SARS-CoV-2 is able to directly promote NETs release by human neutrophils. For that, neutrophils isolated from the blood of healthy controls were cultured in the presence of different MOIs of viable and inactivated SARS-CoV-2. We found that viable SARS-CoV-2, but not inactivated, increased the release of NETs in an MOI-dependent manner (**Fig. 3, A and B**). Importantly, SARS-CoV-2-induced release of NETs was abrogated by Cl-Amidine, PAD-4 inhibitor (**Fig. 3, A and B**).

**Figure 3.**
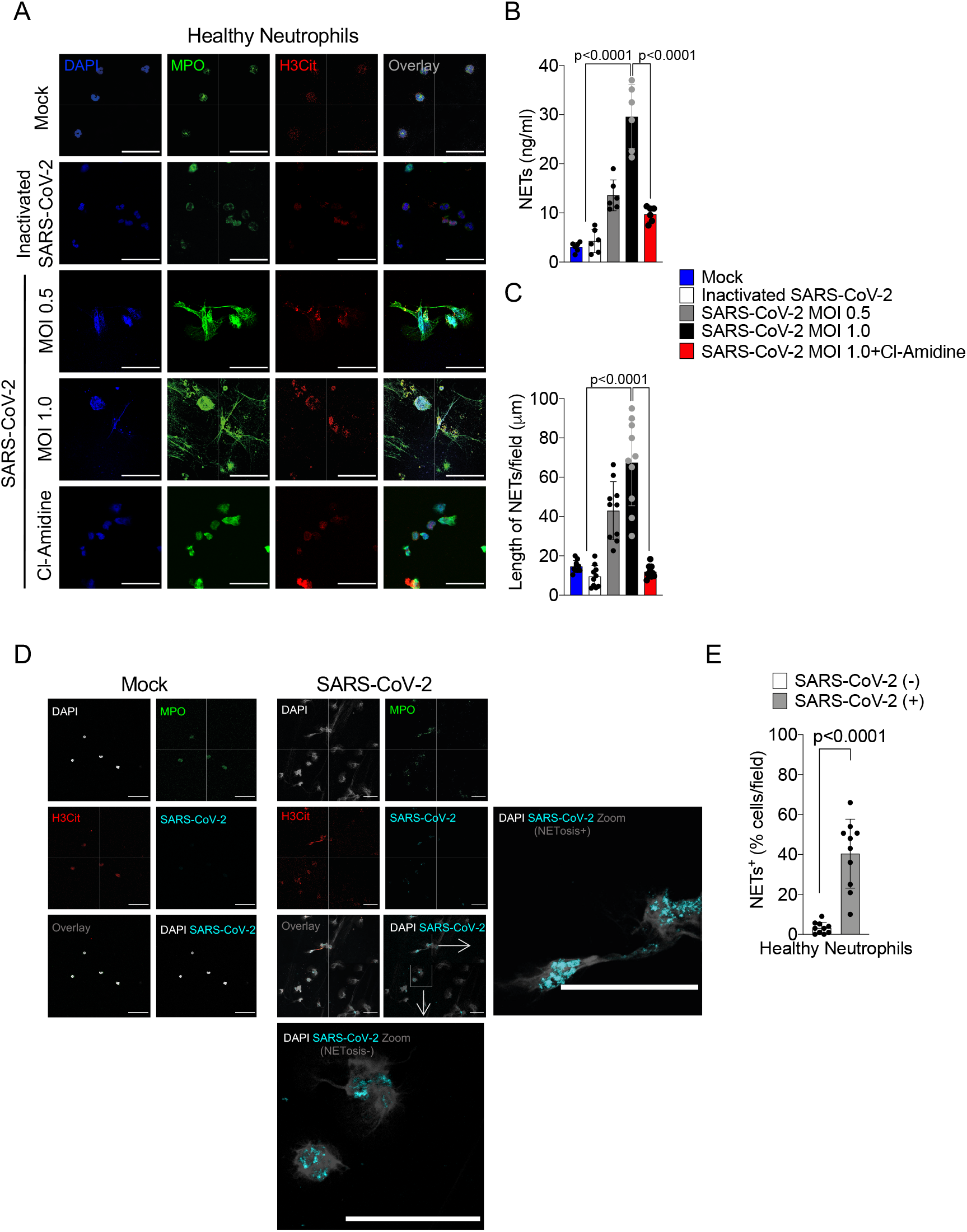
SARS-CoV-2 induces the release of NETs by healthy neutrophils. Neutrophils were isolated from healthy controls and incubated with Mock, inactivated SARS-CoV-2 or SARS-CoV-2 (MOI 0.5 or 1.0). One group of cells incubated with SARS-CoV-2 MOI 1.0 was pretreated with a PAD-4 inhibitor (Cl-Amidine, 200 μM). (A) representative images of NETs release. Cells were stained for nuclei (DAPI, blue), MPO (green), and H3Cit (red). Scale bar indicates 50 μm. (B) NETs quantification by MPO-DNA PicoGreen assay in these neutrophils supernatants. Data are the mean ± SD, one-way ANOVA followed by Bonferroni’s post hoc test. (C) Quantification of NETs length. Data are the mean ± SD, one-way ANOVA followed by Bonferroni’s post hoc test. (D) Representative images showing immunostaining for nuclei (DAPI, white), MPO (green), H3Cit (red), and SARS-CoV-2 (cyan) in neutrophils incubated with Mock or SARS-CoV-2 (MOI 1.0). Scale bar indicates 50 μm. (E) Percentage of NETs positive cells stained, or not, for SARS-CoV-2 antigens. Data are the mean ± SD of 10 fields, two-tailed unpaired Student t-test.

Similar to observed with neutrophils from COVID-19 patients, NETs released by in *vitro* cultured healthy neutrophils in the presence of SARS-CoV-2 have longer branch lengths (**Fig. 3 C**). Notably, we detected neutrophils with SARS-CoV-2 antigens (**Fig. 3 D**). Neutrophils positive for SARS-CoV-2 antigens undergo higher NETosis than negative neutrophils (**Fig. 3, D and E**). SARS-CoV-2 antigens were also detected in the blood neutrophils isolated in the samples from 5 out of 11 COVID-19 patients, and these SARS-CoV-2^+^ neutrophils are also more efficient to produce NETs (**Fig. S3**). These results indicate that SARS-CoV-2 might directly activate neutrophils to release NETs. The lack of NETs release in the culture with inactivated SARS-CoV-2 indicates that binding to the cell receptor is not sufficient to activate the NETs release pathway. This might suggest that active viral infection and/or replication is necessary to trigger NETs release. However, considering that during severe COVID-19, the levels of cytokines and chemokines are increased (Gong et al., 2020), we can not discard that cytokines/chemokines are also activating NETosis during the ongoing COVID-19.

### SARS-CoV-2 activated neutrophils induce lung epithelial cell death through the release of NETs

The release of NETs has been associated with tissue damage in several immune-mediated diseases, including chronic obstructive pulmonary disease, rheumatoid arthritis, and sepsis (Dicker et al., 2018; Khandpur et al., 2013; Colón et al., 2019). NETs cause tissue damage by extracellular exposure of DNA, granular proteins such as MPO and histone, inducing apoptosis and fibrosis processes (Brinkmann et al., 2004). COVID-19 is characterized by extensive damage in the alveolar epithelium (Dolhnikoff et al., 2020). In order to understand the possible role of the released NETs in the pathophysiology of COVID-19, we next explored the hypothesis that NETs would be involved in the damage of lung epithelial cells. To this end, blood isolated neutrophils from healthy controls incubated with SARS-CoV-2 (MOI 1.0) were co-cultured with A549 cells, a human alveolar basal epithelial cell, and cells viability was determined (annexin V^+^ cells) by flow cytometry (**Fig. 4 A**).

**Figure 4.**
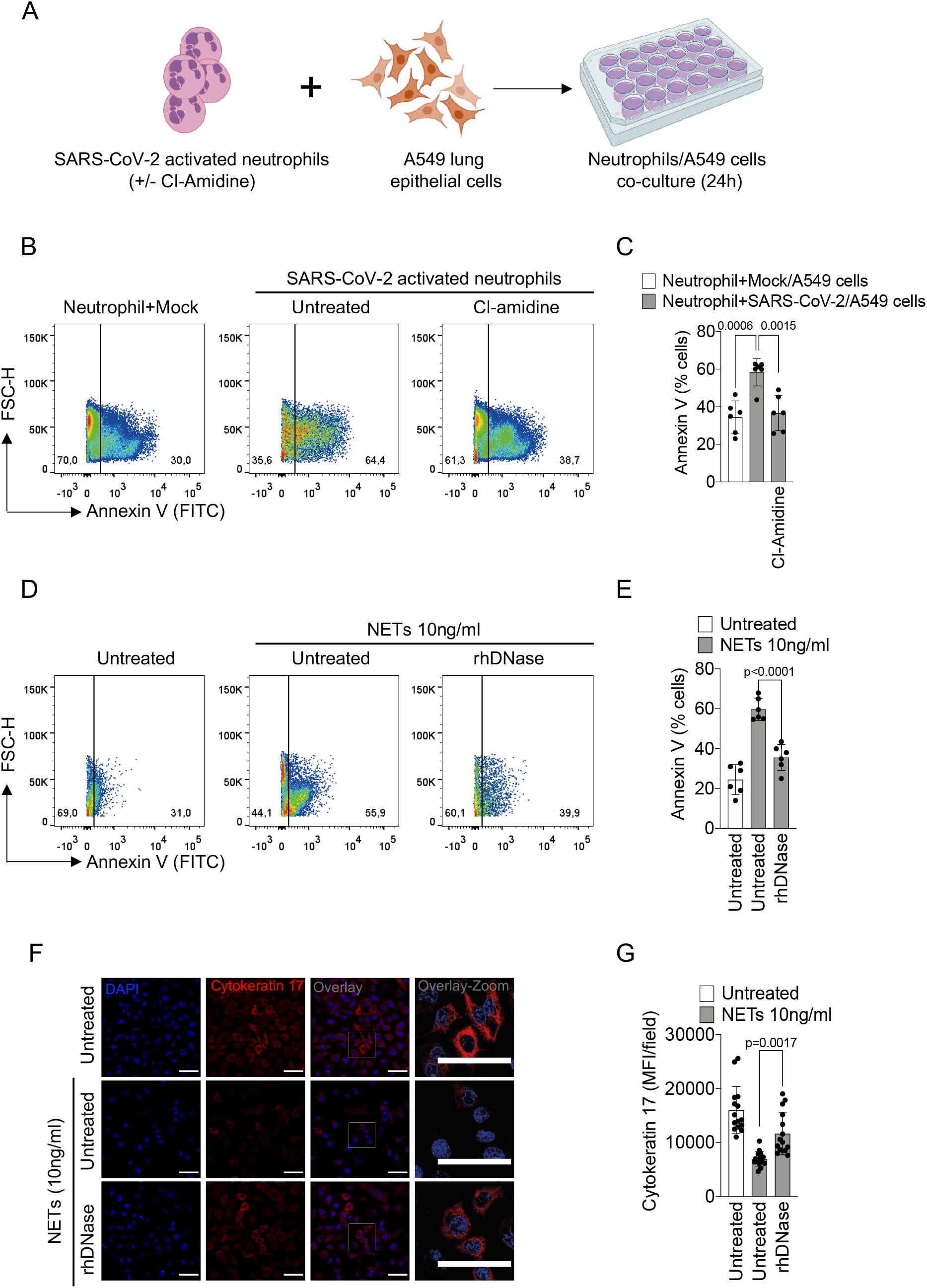
NETs induce the apoptosis of lung epithelial cells. (A) Blood isolated neutrophils (1 × 10^6^ cells) from healthy donors, pretreated, or not, with Cl-Amidine (200 μM) were incubated, or not, with viable SARS-CoV-2 (n=6). After 2 h, these neutrophils were co-cultured with A549 lung epithelial cells (5 × 10^4^ cells) for 24h at 37°C. (B) Representative dot plots of FACS analysis for Annexin V + cells (C) Frequency of Annexin V^+^ A549 cells. Data are the mean ± SD, one-way ANOVA followed by Bonferroni’s post hoc test. (D) NETs were purified from healthy neutrophils stimulated with PMA (50 nM) for 4 h at 37°C. Representative dot plots of FACS analysis of Annexin V+ A549 cells incubated with purified NETs (10 ng/mL) pretreated, or not, with rhDNase (0.5 mg/mL) for 24h at 37°C. (E) Frequency of Annexin V^+^ A549 cells. Data are the mean ± SD. (F) Immunofluorescence analysis of cytokeratin-17 expression in A549 cells incubated for 24 h with purified NETs (10 ng/mL). Cells were stained for nuclei (DAPI, blue) and cytokeratin-17 (red), an epithelial marker. Scale bar indicates 50 μm. (G) Quantification of mean fluorescence intensity of cytokeratin-17 in A549 cells. Data are the mean ± SD of 15 fields/group, one-way ANOVA followed by Bonferroni’s post hoc test.

We found that SARS-CoV-2-activated neutrophils increased the apoptosis of A549 cells compared to non-activated neutrophils. The apoptosis was prevented by treatment of the co-culture with Cl-Amidine (**Fig. 4 B**). Besides releasing higher levels of NETs (**Fig. 1 B**), neutrophils isolated from COVID-19 patients also showed a higher cytotoxic effect on A549 cells than that of neutrophils isolated from healthy controls (**Fig. S4**).

To confirm the deleterious effect of NETs on lung epithelial cells, we evaluate whether NETs purified from *in vitro* Phorbol 12-myristate 13-acetate (PMA)-activated neutrophils (from healthy controls) are also able to cause lung epithelial cells death. We found that the exposure of A549 cells to purified NETs significantly increased the percentage of apoptotic annexin V^+^ cells compared to untreated cells (**Fig. 4, D and E**). Importantly, the addition of rhDNase, which degrades NETs, prevented NETs-induced A549 cell apoptosis to the similar levels observed in untreated cells (**Fig. 4, D and E**). Down-regulation of cytokeratin-17 expression has been associated with reduced viability of epithelial cells (Mikami et al., 2017). The expression of cytokeratin-17 was substantially reduced in A549 cells by the exposure to purified NETs and prevented by the treatment with rhDNase (**Fig. 4, F and G**). Together, these results indicate that NETs might be a potential harmful mediator from neutrophils that cause lung epithelial cell damage during SARS-CoV-2 activation. Moreover, NETs could also activate different PRR receptors, including TLR4 and 7, that mediate the release of inflammatory mediators, which in turn can amplify the direct effects of NETs in COVID-19 patients (Hiroki et al., 2020; Funchal et al., 2015; Saitoh et al., 2012). In this context, during severe COVID-19, it was observed apoptosis of lung epithelial and endothelial cells, events that compromise the lung function, worsening the severity of the disease (Dolhnikoff et al., 2020; Zhang et al., 2020a). In this line, it was observed in the present study the presence of neutrophils releasing NETs, as well as a high concentration of NETs in the tracheal aspirate of COVID-19 patients. Finally, also supporting these findings, it was observed in sections of lung tissues obtained by autopsies of COVID-19 patients, the presence of neutrophils releasing NETs, located in the alveolar space.

The thrombogenic events might contribute to damage to the microcirculation of lungs, hearts, and kidneys in severe COVID-19 patients (Magro et al., 2020; Dolhnikoff et al., 2020; Su et al., 2020; Zhang et al., 2020b). In this context, NETs have been linked with initiation and also enhancement of thrombogenic events in different diseases, directly or through activation of platelets (Perdomo et al., 2019). Although we did not address the possible involvement of NETs in the thrombogenesis during COVID-19, the fact that a systemic increase of NETs production was observed makes it plausible to infer that NETs also trigger these events.

Taking together, our findings demonstrate the potentially deleterious role of NETs in the pathophysiology of severe COVID-19. In conclusion, they support the use of inhibitors of NETs synthesis or promoters of NETs fragmentation, as a strategy to ameliorate the multi-organ damage during the clinical course.

## Materials and methods

### Patients

We enrolled 17 critical and 15 severe patients with COVID-19, according to the Chinese Center for Disease Control and Prevention (10), confirmed by RT-PCR, as described previously (11), or specific antibodies IgM and IgG for SARS-CoV-2 (Asan Easy Test® COVID-19 IgM/IgG kits, Asan Pharmaceutical Co.). Supplementary Table 1 summarizes clinical, laboratory, and treatment records. We performed computed tomography (CT) of the chest for all patients. For comparisons of NETs assays in blood and tracheal aspirate, we collected samples from 21 age and gender-matched healthy controls.

### Plasma and neutrophils isolation

Peripheral blood samples were collected from patients and healthy control by venipuncture and centrifuged at 450*g* for plasma separation. The blood cells were then resuspended in Hank’s (Corning, cat. 21-022-CV) and the neutrophil population was isolated by Percoll (GE Healthcare, cat. 17-5445-01) density gradient. Isolated neutrophils were resuspended in RPMI 1640 (Corning, cat. 15-040-CVR) supplemented with 0.1% BSA The neutrophil purity was >95% as determined by Rosenfeld-colored Cytospin (Laborclin, cat. 620529).

### Tracheal aspirate

The tracheal fluid was obtained by aspiration of the orotracheal tube using an aseptic technique. The collected fluids were mixed 1:1 with 0.1 M Dithiothreitol (DTT, Thermo Fisher Scientific, cat. R0862) and incubated for 15 minutes at 37°C. The solution obtained was centrifuged 750g at 4 °C for 10 min. The cells were pelletized in coverslips with poly-L-lysine solution 0.1% for immunostaining, and the supernatants were used for measurement of NETs. To stimulate airway fluid production and collection from healthy controls, inhalation of 5 mL of hypertonic saline (3%) was performed.

### Virus stock production

The SARS-CoV-2 Brazil/SPBR-02/2020 strain was used for in vitro experiments and was initially isolated from the first Brazilian cases of COVID-19. Stocks were amplified in Vero E6 cell line monolayers maintained in Dulbecco’s Modified Eagle’s (DMEM, Corning, cat. 15-013-CVR). Titers were expressed as the 50% tissue culture infectious dose (TCID50), determined by the Reed-Muench method, and plotted in TCID50 per volume inoculated. For experiments using an inactivated virus, stocks were incubated with formaldehyde at a final concentration of 0.2% overnight at 37°C.

### Production of NETs by isolated neutrophils

Neutrophils (1×10^6^ cells) obtained from COVID-19 patients or healthy controls were incubated with RPMI 1640 supplemented with 0.1% BSA treated or not with Cl-Amidine (200 μM, Sigma-Aldrich, cat. 506282) for 4 h at 37°C. In another context, neutrophils from healthy controls were incubated with SARS-CoV-2 (MOI= 0.5 and 1.0) for 4 h at 37°C or inactivated SARS-CoV-2. The concentration of NETs in supernatants was determined. A total of 5×10^4^ isolated neutrophils were attached to coverslips coated with poly-L-lysine solution 0.1% (Sigma-Aldrich, cat. P8920) incubated for 4 h at 37°C for NETs immunostaining.

### Quantification of NETs

Briefly, plasma or supernatant from neutrophils culture was incubated overnight in a plate pre-coated with anti-MPO antibody (Thermo Fisher Scientific, cat. PA5-16672). The DNA bound to MPO was quantified using the Quant-iT™ PicoGreen® kit (Invitrogen, cat. P11496), as described (5).

### Immunostaining and confocal microscopy

Samples were fixed and stained with the following antibodies: rabbit anti-histone H3 (H3Cit, Abcam, cat. ab5103, 1:500), mouse anti-myeloperoxidase (MPO, 2C7, Abcam, cat. ab25989, 1:500), rabbit anti-Cytokeratin 17 (Abcam, ab53707, 1:400). The samples were washed in PBS and incubated with secondary antibodies Donkey anti-Mouse IgG AlexaFluor 647 (Thermo Fisher Scientific, cat. A32787, 1:800) or AlexaFluor 488 (Abcam, cat. ab150061, 1:800) and Donkey anti-rabbit IgG AlexaFluor 488 (Abcam, cat. ab150065, 1:800) or AlexaFluor 594 (Abcam, cat. ab150076, 1:800). The nuclei were stained with 4’,6-Diamidino-2-Phenylindole, Dihydrochloride (DAPI, Life Technologies, cat. D1306, 1:1,000). To detect SARS-CoV-2 for immunostaining, we used a human serum kindly provided by Dr. Edison Durigon, from a recovered COVID-19 patient (1:400). We used anti-human IgG biotin-conjugated (Sigma-Aldrich, cat. B-1140, 1:1,000) followed by amplification kit TSA Cyanine 3 System (Perkin Elmer, cat. NEL704A001KT), according to the manufacturer’s protocol. Images were acquired by Axio Observer combined with LSM 780 confocal device with 630 x magnification (Carl Zeiss). The data were performed with Fiji/ImageJ software.

### Quantification of NETs length

The immunostained images acquired were analysed to examine the length of NETs. DAPI, MPO, and H3Cit staining were selected to identify pixels present in the NETs. We used the adapted plugin Simple Neurite Tracer from Fiji by ImageJ.

### Purification of NETs

Isolated neutrophils (1.5×10^7^ cells) from healthy controls were stimulated with 50 nM of Phorbol 12-myristate 13-acetate (PMA, Sigma-Aldrich, cat. P8139) for 4 h at 37°C. The medium containing the NETs was centrifuged at 450 g to remove cellular debris, and NETs-containing supernatants were collected and centrifuged at 18,000g. Supernatants were removed, and pellets were resuspended in PBS. NETs were then quantified through GeneQuant (Amersham Biosciences Corporation, Piscataway).

### Epithelial cell damage assay

Human alveolar basal epithelial A549 cell line (5×10^4^), maintained in DMEM, were co-cultured with Cl-Amidine (200μM) pre-incubated (30 min) neutrophils from healthy controls (1×10^6^) and SARS-CoV-2 (MOI 1.0) for 2 h. The A549 cells were also co-cultured with neutrophils from COVID-19 patients or with purified NETs (10ng/mL) pretreated, or not, with rhDNase (0.5 mg/mL, Roche, 2h at 37°C). The co-cultures of A549 cells with neutrophils or NETs were incubated 24h at 37°C, and the A549 viability was determined by flow cytometric analysis of Annexin V staining or immunostaining analysis for cytokeratin-17.

### Flow cytometry

Briefly, whole blood leukocytes were stained with Fixable Viability Dye eFluor™ 780 (eBioscience, cat. 65-0865-14, 1:3,000) and monoclonal antibodies specific for CD14 (M5E2, BD, cat. 557153, 1:50), CD19 (HIB19, Biolegend, cat. 302212, 1:200), CD15 (W6D3, BD, cat. 563141, 1:200) and CD16 (ebioCB16(CB16), eBioscience, cat. 12-0168-42, 1:200) for 30 min at 4°C. A549 cells (5×10^4^) were stained with FITC ApoScreen® Annexin V Apoptosis Kit (SouthernBiotech, cat. 10010-02), according to the manufacturer’s instructions. All data were collected on FACSVerse flow cytometers (BD Biosciences) for further analysis using FlowJo (TreeStar) software.

### Lung samples from autopsies

Ten COVID-19 patients were autopsied with the ultrasound-guided minimally invasive approach (13). The COVID19-HC-FMUSP autopsy study was approved by the HC-FMUSP Ethical Committee (protocol #3951.904) and performed at the “ Image Platform in the Autopsy Room” (https://pisa.hc.fm.usp.br/). The sampling protocol was previously described (Duarte-Neto et al., 2019). Pulmonary tissue samples were stained with hematoxylin and eosin (H&E) and immunostaining.

### Statistics

Statistical significance was determined by either two-tailed paired or unpaired Student t-test and one-way ANOVA followed by Bonferroni’s post hoc test; p<0.05 was considered statistically significant. Statistical analyses and graph plots were performed with GraphPad Prism 8.4.2 software.

### Study approval

The procedures followed in the study were approved by the National Ethics Committee, Brazil (CONEP, CAAE: 30248420.9.0000.5440). Written informed consent was obtained from recruited patients.

## Data Availability

The data supporting the findings of this study are available within the article [and/or] its supplementary materials.

## Author Contributions

F.P.V., M.C.P., F.Q.C., R.D.R.O., J.C.A.F., T.M.C., P.L.J., E.A., L.O.L and L.D.C. contributed to the study design. J.E.T-K., M.H.F.L., D.C.N. and D.F.C. performed neutrophils isolation. F.P.V., D.B.C., R.R.C.R and G.A. performed immunostaining and confocal analysis. J.E.T-K., A.H.S. and D.B.C. performed NETs quantification. C.M.S. performed tracheal fluid experiments. T.M., M.D., A.N.D-N. and P.H.N.S. contributed to lung autopsies analysis. F.P.V. and M.C.P. performed A549 epithelial cell damage assay. A.H.S. performed the purification of NETs. F.P.V., M.C.P., R.B.M and I.A.C. performed SARS-CoV-2 experiments. R.B.M. performed RT-PCR and IgM/IgG assay for SARS-CoV-2. F.P.V., M.H.F.L. and D.C.N. performed FACS analysis. M.I.F.L., M.N.B., L.P.B., M.C.G., R.L-A., S.C.L.A., F.C.V., R.C.S., V.R.B., M.A.M., C.H.M., M.C.B., A.P.F., F.D-P. and A.T.F. contributed to the collection of clinical specimens, demographic and clinical characteristics analysis from COVID-19 patients. F.Q.C., R.D.R.O., T.M.C., F.P.V., J.C.A.F., M.C.P., P.L.J., E.A., D.S.Z. and L.O.L. wrote the manuscript. All authors approved the manuscript.

## Acknowledgments

We are grateful to Marcella Daruge Grando, Livia Maria C. S. Ambrósio, Muriel C. R. O. Berti, Basílica Botelho Muniz and Juliana Trench Abumansur for technical assistance.

## Supplemental Figures Legends

**Supplementary Table 1 – Demographic and clinical characteristics of COVID-19 patients**.

**Figure S1.**
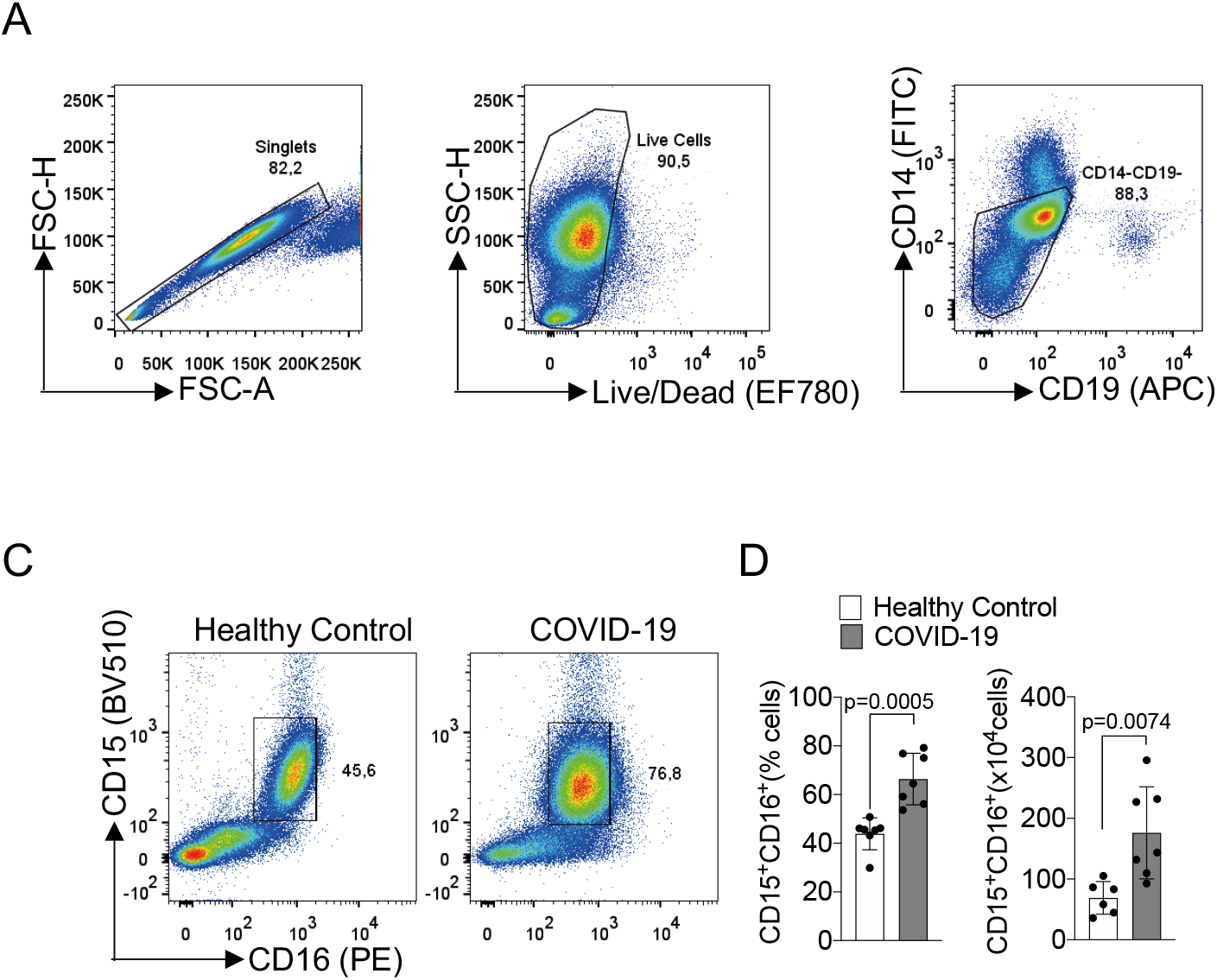
Representative gating strategies used for flow cytometry analysis of CD15^+^CD16^+^ neutrophils from whole blood. (A) Doublet cells were excluded by FSC-H and FSC-A gating for all flow cytometry analysis. Viable cells were identified using fixable viability stain and SSC-A gating. Neutrophils were identified as cells stained for CD15^+^CD16^+^ among CD14^-^CD19^-^ cells. (B) Flow cytometry analyses of living cells from the whole blood of healthy controls or COVID-19 patients. (c) Frequency and absolute numbers of CD15^+^CD16^+^ neutrophils gated on CD14^-^CD19^-^ live cells from whole blood from healthy controls (n=7) or COVID-19 patients (n=7). Data are the mean ± SD, two-tailed unpaired Student t-test.

**Figure S2.**
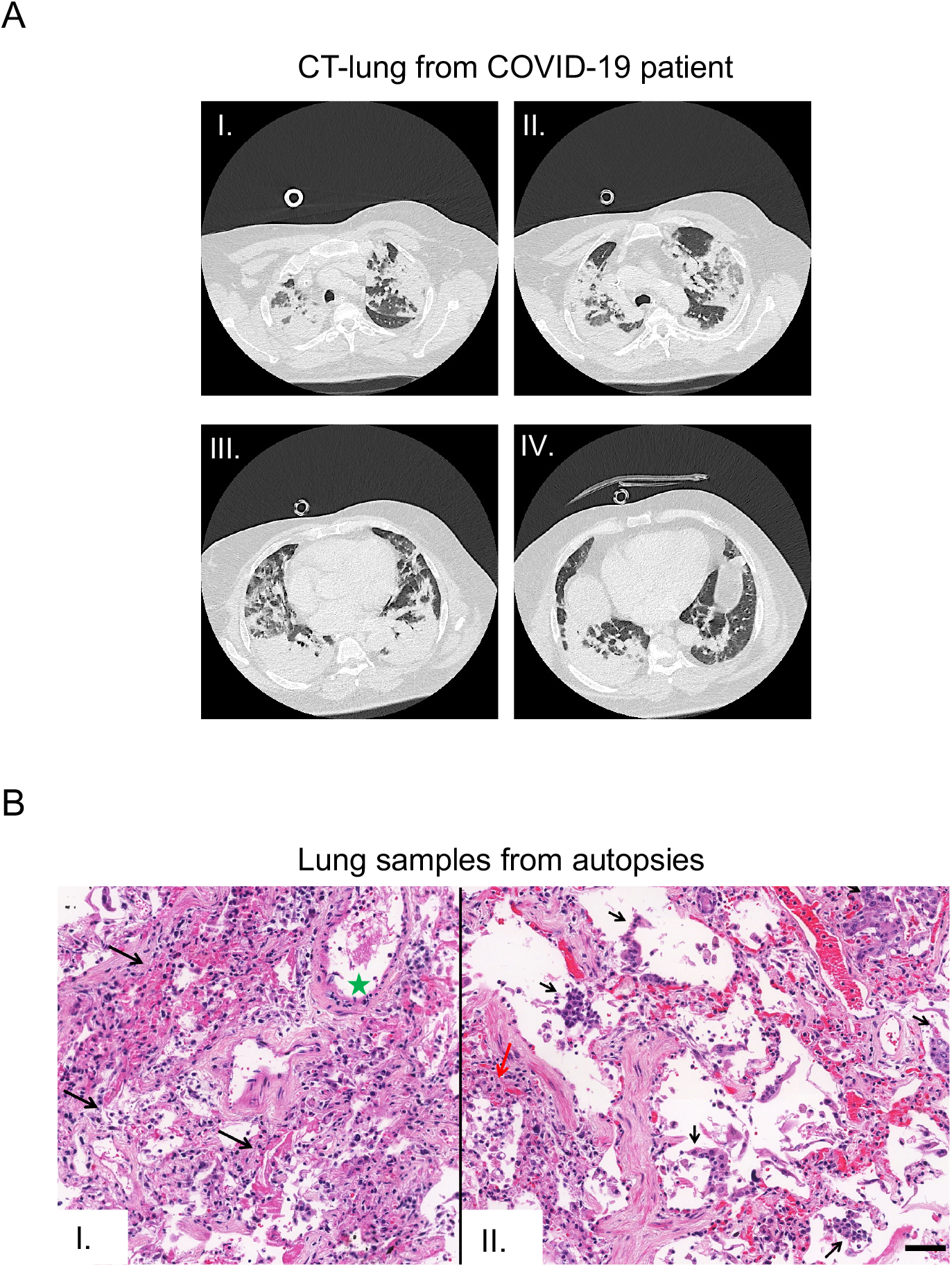
Lung histopathological analysis in fatal cases of COVID-19. (A) Computed tomography (CT) of the chest of one patient who died from COVID-19. Images from apical to basal segments (I to IV) show multiple consolidations with air bronchograms in a peripheral and peribronchovascular distribution, more evident in the lower lobes, associated with ground-glass opacities. (B) Representative pulmonary histological findings in ten cases, autopsied by ultrasound guided-minimally invasive autopsy. I. The area with interstitial and alveolar neutrophilic pneumonia with diffuse alveolar damage and hyaline membranes in the alveolar space (black arrows). Septal vessel with margination of leukocytes and an intraluminal early fibrin thrombus (green star). II, Area with neutrophilic pneumonia (red arrow), septal thickening, epithelial desquamation, and squamous metaplasia (black arrows). (I and II: H&E). Scale bar indicates 50 μm.

**Figure S3.**
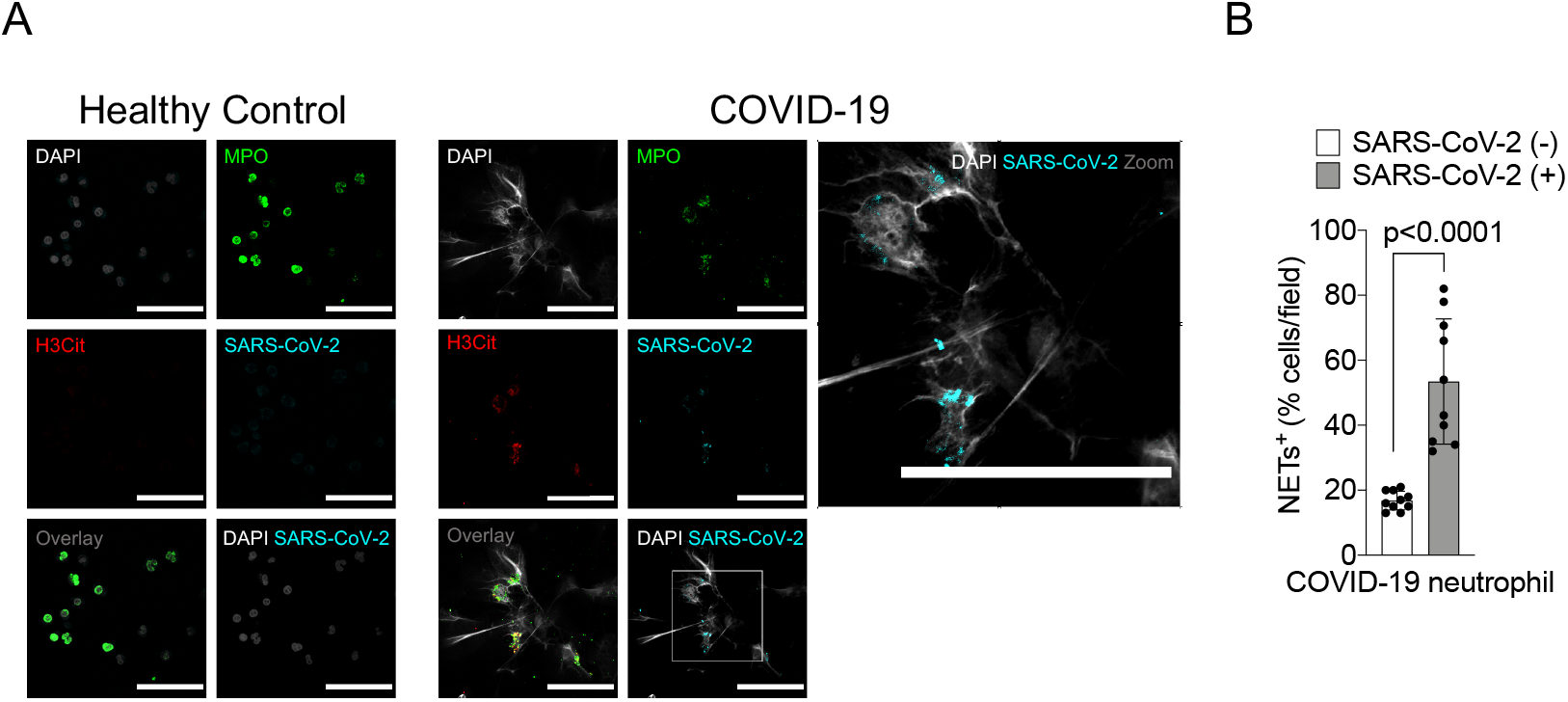
Detection of SARS-CoV-2 antigens in blood neutrophils from COVID-19 patients. (A) Representative confocal images showing the detection of SARS-CoV-2 antigens in blood neutrophils from COVID-19 patients (n=3), but not in neutrophils from healthy controls (n=3). Cells were stained for nuclei (DAPI, white), MPO (green), H3Cit (red), and SARS-CoV-2 (Cyan). Scale bar indicates 50 μm. (B) Percentage of NETs positive cells stained, or not, for SARS-CoV-2 antigens. Data are the mean ± SD of 5 fields/group, two-tailed unpaired Student t-test.

**Figure S4.**
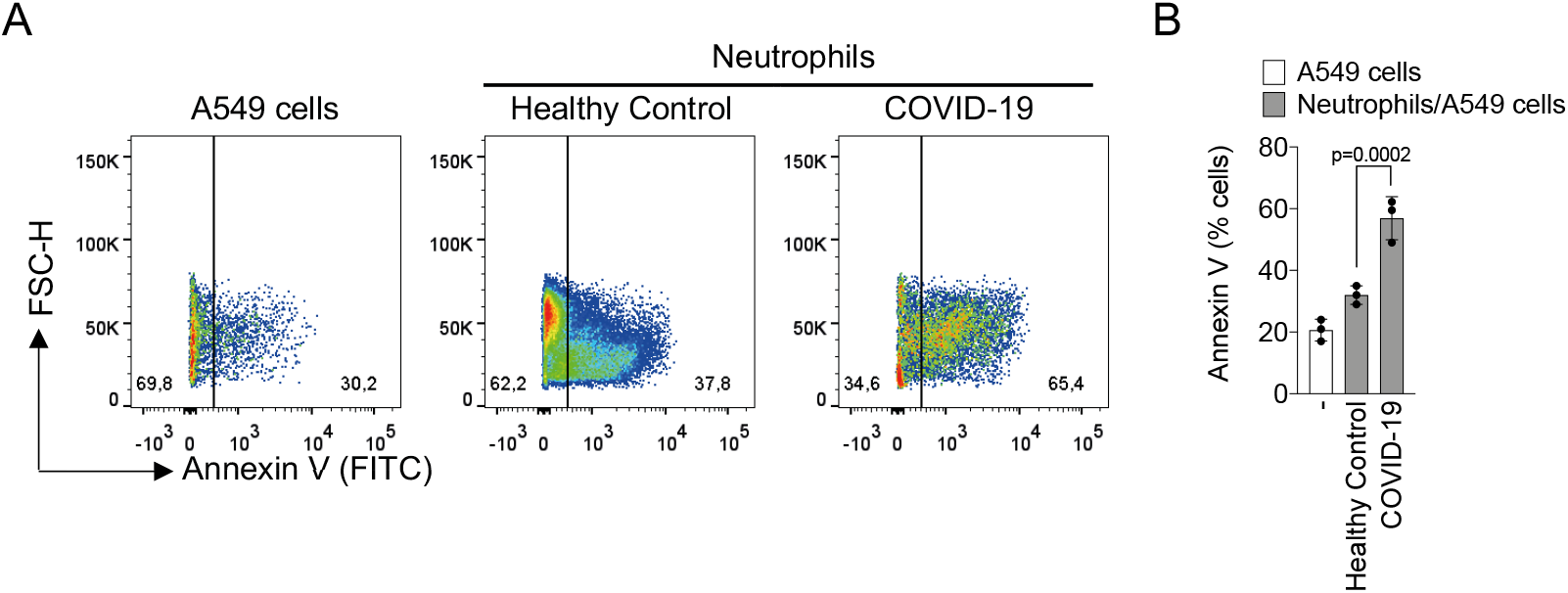
Blood-isolated neutrophils from COVID-19 patients induce lung epithelial cell apoptosis. Blood isolated neutrophils (1×106 cells) from healthy controls (n=3) or COVID-19 patients (n=3) were co-cultured with A549 lung epithelial cells (5×104 cells) for 24h at 37°C. (A) Representative dot plots of FACS analysis for A549 Annexin V+ cells. (B) Frequency of Annexin V+ A549 cells. Data are the mean ± SD, one-way ANOVA followed by Bonferroni’s post hoc test.

**Table 1:**
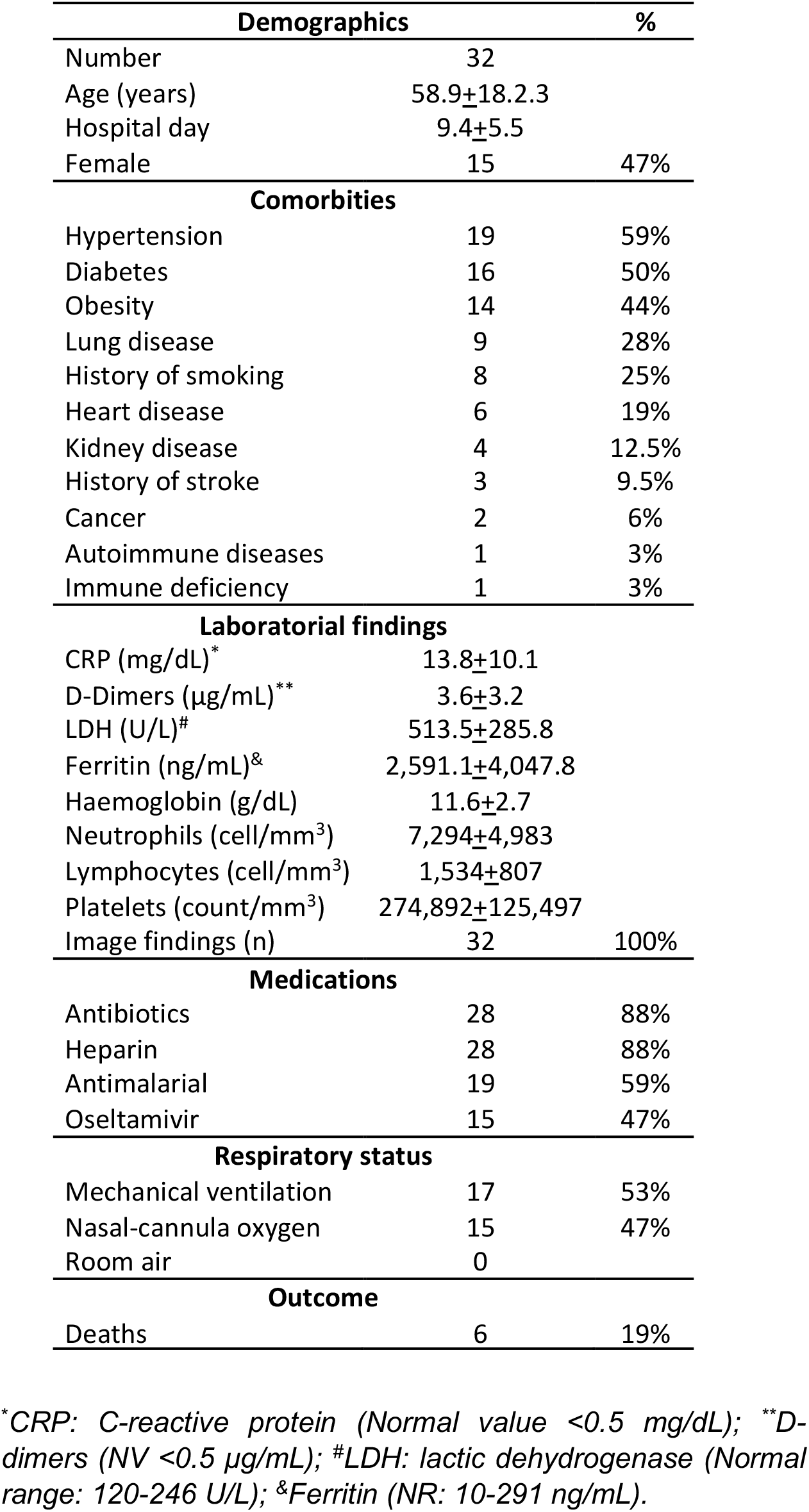
COVID-19 patient characteristics

